# Immune history shapes human antibody responses to H5N1 influenza viruses

**DOI:** 10.1101/2024.10.31.24316514

**Authors:** Tyler A. Garretson, Jiaojiao Liu, Shuk Hang Li, Gabrielle Scher, Jefferson J.S. Santos, Glenn Hogan, Marcos Costa Vieira, Colleen Furey, Reilly K. Atkinson, Naiqing Ye, Jordan Ort, Kangchon Kim, Kevin A. Hernandez, Theresa Eilola, David C. Schultz, Sara Cherry, Sarah Cobey, Scott E. Hensley

## Abstract

Avian H5N1 influenza viruses are circulating widely in cattle and other mammals and pose a risk for a human pandemic. Previous studies suggest that older humans are more resistant to H5N1 infections due to childhood imprinting with other group 1 viruses (H1N1 and H2N2); however, the immunological basis for this is incompletely understood. Here we show that antibody titers to historical and recent H5N1 strains are highest in older individuals and correlate more strongly with year of birth than with age, consistent with immune imprinting. After vaccination with an A/Vietnam/1203/2004 H5N1 vaccine, both younger and older humans produced H5-reactive antibodies to the vaccine strain and to a clade 2.3.4.4b isolate currently circulating in cattle, with higher seroconversion rates in young children who had lower levels of antibodies before vaccination. These studies suggest that younger individuals might benefit more from vaccination than older individuals in the event of an H5N1 pandemic.

## Main text

Highly pathogenic avian influenza (HPAI) clade 2.3.4.4b H5 viruses began circulating at high levels in bird populations across the world in 2020^1-4^ and have caused infections in mammals such as foxes, seals, and mink^5-8^. At the end of 2023, a clade 2.3.4.4b H5N1 virus began circulating in dairy cattle in the United States^9^, with widespread transmission between cows^10^. Clade 2.3.4.4b H5N1 viruses do not currently bind well to receptors found in human upper airways^11,12^; however, widespread circulation in mammals could lead to adaptive substitutions that increase viral attachment, replication, and human transmission^13,14^.

Previously circulating H5N1 viruses caused higher mortality rates in younger humans relative to older humans^15,16^. It is possible that immunity elicited by seasonal influenza viruses affects H5N1 susceptibility. Influenza A viruses can be broadly split into 2 different phylogenetic groups^17^. Group 1 (H1N1 and H2N2) and group 2 (H3N2) viruses have circulated during distinct times since 1918 (**Fig. 1a**), and therefore immunity against each of these viruses is partly shaped by an individual’s birth year (**Extended Fig. 1**). Gostic and colleagues proposed that individuals born before 1968 may be more refractory to severe disease following H5N1 (a group 1 virus) infection since most of these individuals were likely ‘immunologically imprinted’ with other group 1 viruses (H1N1 and H2N2) in childhood^18^. Although H5N1 is antigenically distinct from H1N1 and H2N2, all three of these viruses share homology in conserved epitopes, including the hemagglutinin (HA) stalk domain^17^. Previous studies reported that H1 stalk-reactive antibodies are more prevalent in older individuals^19,20^; however, a subsequent study found no clear association with H1 stalk-reactive antibodies and birth year among adults^21^.

**Fig. 1.**
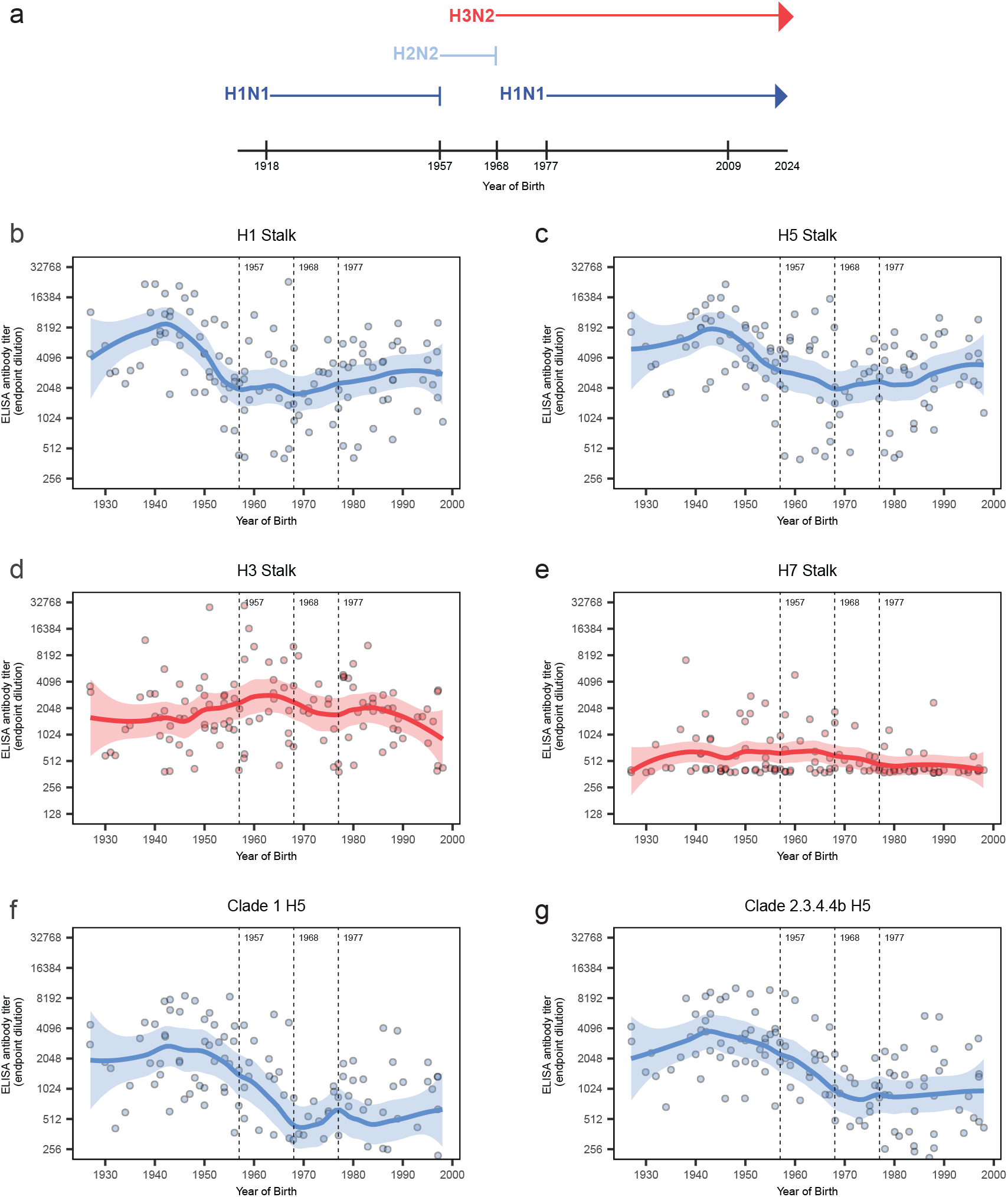
Group 1 immune imprinting primes robust H5-reactive antibody responses. (**a**) Group 1 (blue) and group 2 (red) influenza viruses have circulated at distinct times since 1918. Sera samples were collected from healthy donors (n=121) at the Hospital of the University of Pennsylvania in 2017 and we quantified IgG binding in ELISA to ‘headless’ A/California/4/2009 H1 stalk (**b**), ‘headless’ A/Vietnam/1203/2004 H5 stalk (**c**), ‘headless’ A/Finland/486/2004 H3 stalk (**d**), A/Shanghai/02/2013 H7 stalk (**e**), a clade 1 A/Vietnam/1203/2004 full length H5 HA (**f**) and a clade 2.3.4.4b A/Pheasant/New York/22-009066-011/2022 full length H5 HA (**g**). (**b-g**) Vertical dashed lines mark years of the 1957 H2N2 and 1968 H3N2 pandemics and 1977 reemergence of H1N1. Each circle represents a geometric mean antibody titer in serum from a single donor from two independent replicates, and the trend lines are locally estimated scatterplot smoothing (LOESS) curves (smoothing parameter = 0.4, degree = 2) with 95% confidence intervals.

We quantified antibodies reactive to group 1 HA stalks (H1 and H5) and group 2 HA stalks (H3 and H7) in serum samples collected in 2017 from 121 healthy adults born between 1927-1998 (19-90 years old at time of sampling). H1 and H5 stalk-reactive antibodies were higher in older adults, although younger adults possessed moderate amounts of these antibodies (**Fig. 1b-c**). We asked if stalk-reactive antibody levels correlated with the probability of immune imprinting (i.e., initial infection in childhood) with viruses of the same subtype or group, estimated from historical data. H1 and H5 stalk-reactive antibody titers were both positively correlated with group 1 and H1N1 imprinting probabilities (**Table S1**). We found that group 2 HA stalk antibody levels were generally lower with no obvious correlation between titer and birth year (**Fig. 1d-e, Table S1**). H3 stalk-reactive antibodies were present in sera at similar levels regardless of donor age (**Fig. 1d**) and H7 stalk-reactive antibodies were generally low in the sera from most donors (**Fig. 1e**).

Consistent with the group 1 HA stalk titer data, we found that individuals imprinted with group 1 viruses in childhood possessed high levels of antibodies reactive to clade 1 H5 full length HA (A/Vietnam/1203/2004) (**Fig. 1f, Table S1**) and clade 2.3.4.4b H5 full length HAs (A/Pheasant/New York/22-00906-001/2022 and A/Dairy Cattle/Texas/24-008749-002-v/2024) (**Fig. 1g, Extended Data Fig. 2**, and **Table S1**). Most of these cross-reactive antibodies were non-neutralizing, although we detected antibodies that neutralized clade 1 and clade 2.3.4.4b H5N1 in rare individuals of all ages (**Extended Data Fig. 3**). Taken together, our data suggest that childhood exposures to H1N1 and H2N2 prime mostly non-neutralizing group 1 HA stalk antibodies that bind to diverse H5 HAs.

**Fig. 2.**
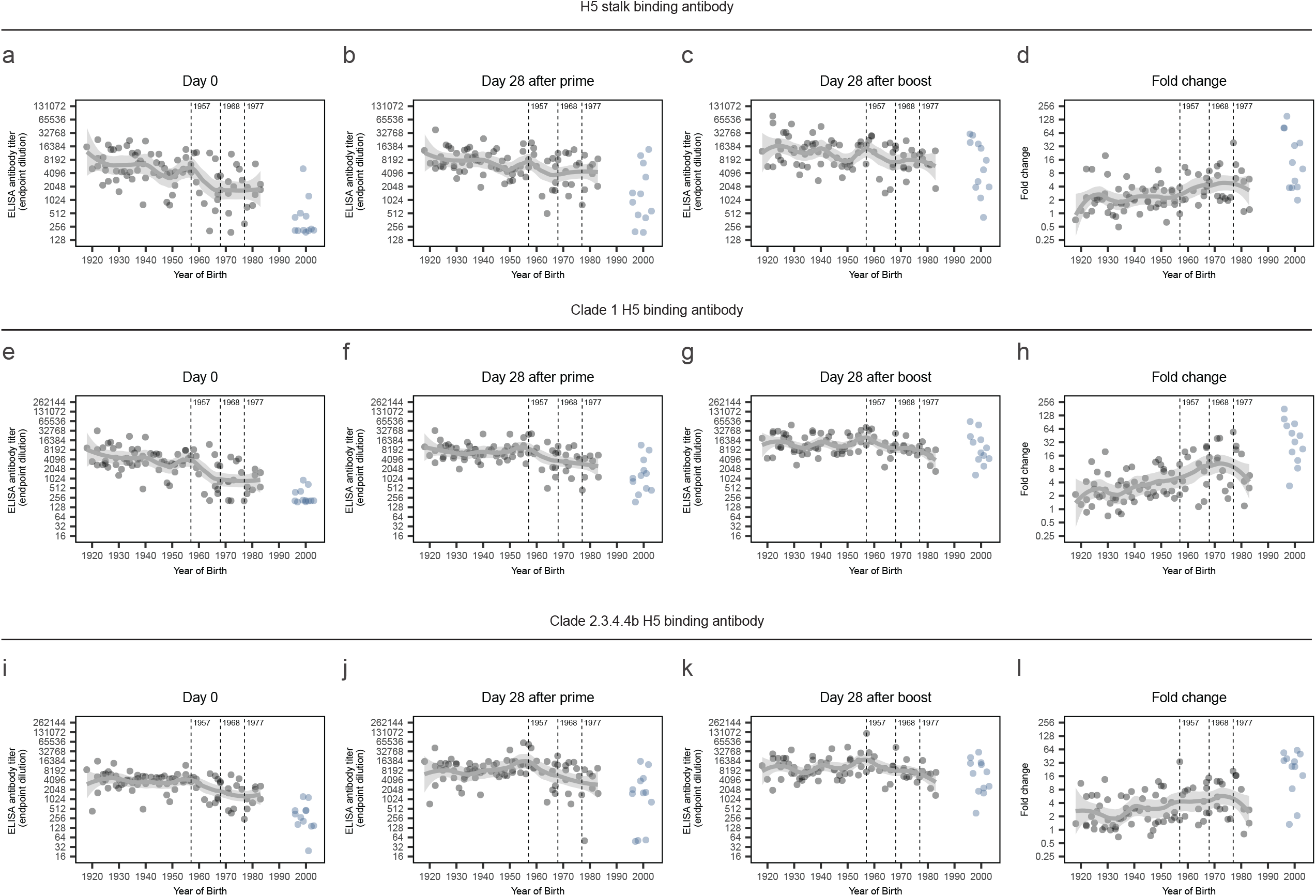
H5N1 vaccination elicits strong H5 stalk-reactive antibodies in children. Sera samples were obtained from participants (n=100) before (day 0) and 28 days after receiving a first and second dose of an unadjuvanted A/Vietnam/1203/2004 H5N1 vaccine in 2005-2006. We quantified IgG binding to a ‘headless’ A/Vietnam/1203/2004 H5 stalk (**a-c**), and we calculated titer fold change by dividing day 28 post-boost titers by day 0 titers (**d**). We also measured antibody binding and fold change to clade 1 A/Vietnam/1203/2004 full length H5 (**e-h**) and clade 2.3.4.4b A/Pheasant/New York/22-009066-011/2022 full length H5 (**i-l**). Vertical dashed lines mark years of the 1957 H2N2 and 1968 H3N2 pandemics and 1977 reemergence of H1N1. Each circle represents a geometric mean antibody titer in serum from a single donor from two independent replicates. Samples from adults are grey and children are blue. Trend lines are locally estimated scatterplot smoothing (LOESS) curves (smoothing parameter = 0.4, degree = 2) with 95% confidence intervals, fitted to adult samples.

**Fig. 3.**
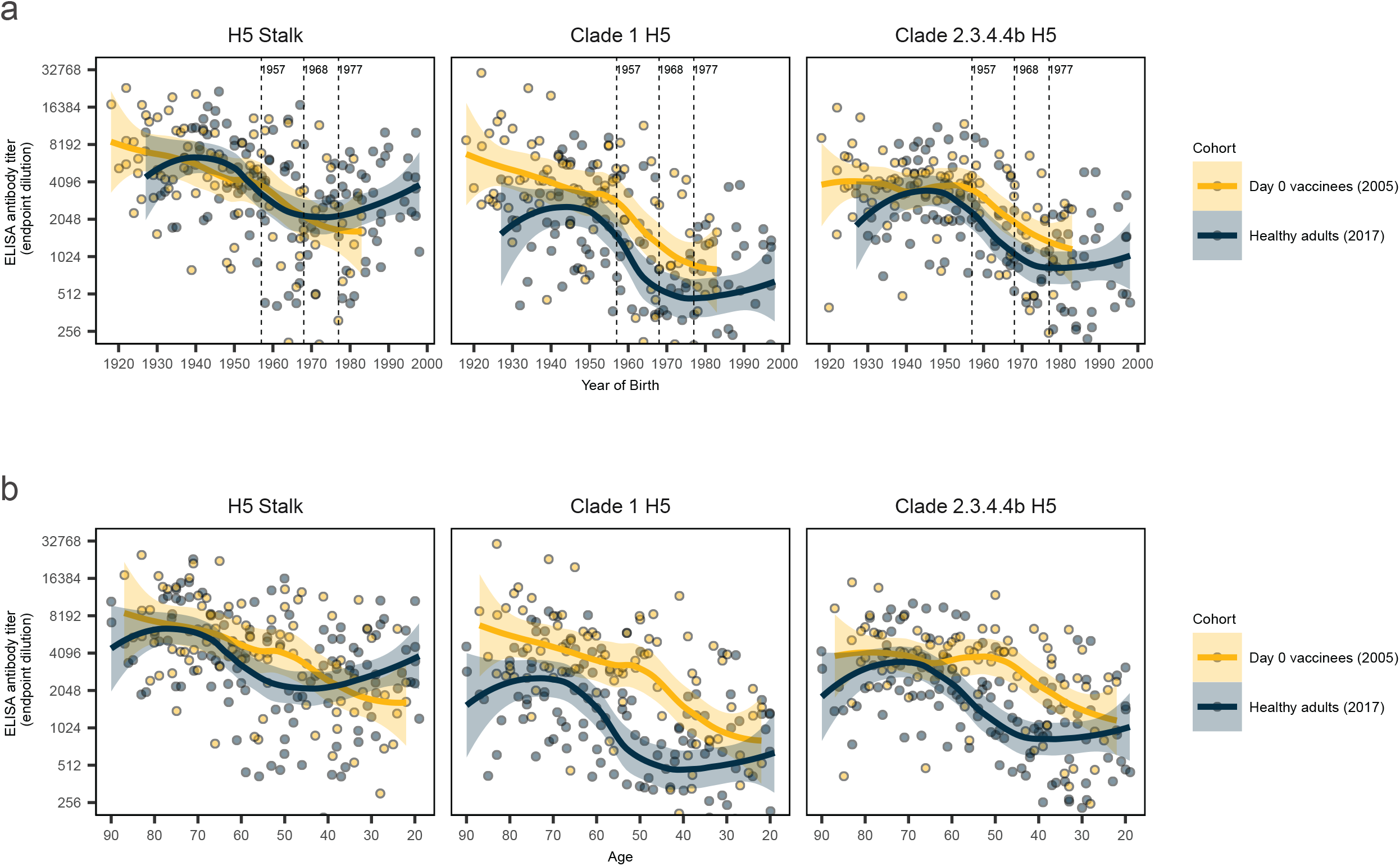
H5-rective antibody levels correlate better with birth year compared to age. Antibody titers in samples collected in 2005 (yellow) and 2017 (blue) against H5 stalk, clade 1 A/Vietnam/1203/2004 full length H5, and clade 2.3.4.4b A/Pheasant/New York/22-009066-011/2022 full length H5 are shown as a function of birth year (**a**) and age (**b**). Vertical dashed lines mark years of the 1957 H2N2 and 1968 H3N2 pandemics and 1977 reemergence of H1N1 (**a**). Each circle represents a geometric mean antibody titer in serum from a single donor from two independent replicates. Trend lines are locally estimated scatterplot smoothing (LOESS) curves (smoothing parameter = 0.65, degree = 2) with 95% confidence intervals, fitted to adult samples.

Previous studies showed that H5 vaccines elicit HA stalk antibodies in humans^22,23^; however, it is unknown if H5 vaccines elicit different levels of HA stalk antibodies in individuals with different birth years. We obtained serum samples collected from children and adults (born between 1918-2003) before and after administration of a 45 µg dose of an H5N1 (A/Vietnam/1203/2004) unadjuvanted vaccine (ClinicalTrials.gov #s: NCT00133536, NCT00230750, NCT00115986^24^). Since these clinical studies were completed in the years 2005-2006, the participants ranged from 2-97 years of age at time of vaccination. All participants received a prime-boost regiment, and we analyzed serum samples before vaccination and 28 days after the 1^st^ and 2^nd^ vaccination.

Consistent with our previous results, older adults possessed high levels of H5 stalk-reactive antibodies before vaccination (**Fig. 2a**). H5 stalk-reactive antibody levels increased slightly in older individuals and more substantially in children after each vaccination (**Fig. 2b-c**). We observed the highest fold change in H5 stalk-reactive antibody titers in children, whose antibody levels were lower prior to vaccination (**Fig. 2d**). We obtained similar data when we measured antibodies reactive to the full length A/Vietnam/1203/2004 HA vaccine antigen (**Fig. 2e-h**). Older individuals possessed higher levels of A/Vietnam/1203/2004 HA-reactive antibodies relative to children before vaccination (**Fig. 2e**) and these antibodies were boosted in participants in all age groups after vaccination (**Fig. 2f-g**) with the greatest benefit in children (**Fig. 2h**). Consistent with a recent report^25^, we found that the clade 1 A/Vietnam/1203/2004 H5N1 vaccine elicited antibodies that bound to the more recent antigenically distinct clade 2.3.4.4b HA (**Fig. 2i-l**). Antibody levels against the clade 1 and clade 2.3.4.4b HAs were similar after vaccination (compare **Fig. 2g** and **Fig. 2k**).

While some HA stalk antibodies are broadly neutralizing, many of these antibodies protect through Fc receptor-dependent mechanisms such as antibody dependent cellular cytotoxicity (ADCC)^26^. We therefore determined if antibodies elicited by the A/Vietnam1203/2004 vaccine neutralized H5 virus or mediated ADCC with matched and mismatched H5 HAs. The vaccine elicited antibodies that neutralized clade 1 virus (**Extended Data Fig. 4a-c**) with larger fold-change titer increases in children (**Extended Data Fig. 4d**). The clade 1 vaccine antigen also elicited clade 2.3.4.4b neutralizing antibodies in some participants, but this was more sporadic (**Extended Data Fig. 4e-h**). ADCC activity against the clade 1 and clade 2.3.4.4b H5N1 viruses was high in serum from older individuals prior to vaccination (**Extended Data Fig. 4i and 4m**) and increased in younger individuals following vaccination (**Extended Data Fig. 4j-k and 4n-o**), with the largest ADCC titer fold change in children following vaccination (**Extended Data Fig. 4l and 4p**).

Finally, we directly compared antibody titers from our healthy donors (**Fig. 1**) with pre-vaccination titers from adults from the H5 vaccine trials (**Fig. 2**). Since these samples were collected 12 years apart (2005 versus 2017), we were able to determine if H5 antibody levels were more closely associated with birth year or age (**Fig. 3a-b**).

Consistent with immune imprinting, antibody levels were similar across these datasets when plotted as a function of birth year (**Fig. 3a**) but different when plotted as a function of age (**Fig. 3b**). Using two different statistical approaches, we found that titers to full length H5 proteins from clade 1 and clade 2.3.4.4b (but not to the H5 stalk) had stronger statistical associations with year of birth and group 1 imprinting probability than with age (**Tables S2** and **S3**). Antibody levels were less strongly associated with H1N1 imprinting than with group 1 imprinting, suggesting that initial infections with H1N1 or H2N2 prime antibody responses against clade 1 and clade 2.3.4.4b H5 proteins. Taken together, these analyses corroborate the role of imprinting with group 1 viruses in shaping antibody levels to H5N1 in humans.

Further studies need to be completed to determine if strong immunological imprinting is solely a byproduct of initial influenza virus encounters or if multiple exposures during childhood are required to prime robust memory B cell responses. Our previous studies suggest that heterosubtypic influenza virus infections in ferrets and humans boost HA stalk responses against the originally infecting viral subtype^27^.

Longitudinal cohort studies, in which infections and vaccinations are tracked carefully from birth, are required to better understand how childhood exposures impact the generation of influenza virus antibodies later in life. In our experiments, younger individuals possessed lower levels of H5-reactive antibodies prior to vaccination, and it is likely that many of the younger children in our study had not yet been exposed to group 1 viruses at the time of sample collection.

Our study has limitations. For our vaccination studies, we analyzed serum samples from 3 separate clinical studies to compare antibody responses elicited in individuals with diverse birth years. The same vaccine formulation, dose, and vaccination schedule were used in all 3 clinical studies, so it is unlikely that the observed differences were due to clinical study differences. We only compared antibody responses elicited by unadjuvanted vaccines in our studies. Khurana and colleagues recently reported that adjuvanted clade 1 H5N1 vaccines elicit antibodies with high reactivity to clade 2.3.4.4b H5N1 viruses^25^, and it will be important for future studies to evaluate if there are birth year-related differences in responses to adjuvanted vaccines. We only evaluated antibody responses to HA, and additional studies should be completed to determine if humans possess cross-reactive antibodies against the neuraminidase protein of H5N1. Finally, all our vaccine studies were based on clade 1 H5N1 antigens, and future studies should evaluate responses to other vaccine antigens based on more contemporary H5N1 strains.

Based on our studies and the observation that H5N1 viruses have typically caused more disease in younger individuals^15,16^, it is possible that older individuals would be partially protected in the event of an H5N1 pandemic. Younger individuals who have fewer group 1 influenza virus exposures would likely benefit more from an H5N1 vaccine, even a mismatch stockpiled vaccine^28^. It will be important to continue to test new updated vaccine antigens in individuals with diverse birth years, including children. It will also be important to closely monitor clade 2.3.4.4b H5N1 virus circulating in wild and domestic animals, as well as spillover infections of humans, so that we can continue to evaluate the pandemic risk of these viruses.

## Data Availability

All data are included in the manuscript and there are no restrictions. The code implementing the analyses is available at https://github.com/cobeylab/H5_titers_vs_imprinting

## Acknowledgments

We thank the Penn Medicine BioBank as a resource for obtaining serum from humans with different birth years. We thank NIAID and the VTEU clinical study teams from DMID 04-063, DMID 04-076, and DMID04-0077 for providing sera samples from clinical studies. This project was funded in part with Federal funds from the National Institute of Allergy and Infectious Diseases, National Institutes of Health, Department of Health and Human Services, under Contract Nos. 75N93021C00015 (S.E.H. and S.C.) and grant numbers R01AI08686 (S.E.H.). S.E.H. holds an Investigators in the Pathogenesis of Infectious Disease Awards from the Burroughs Wellcome Fund.

## Author Contributions Statement

S.E.H., T.A.G., J.L., S.H.L., G.S., C.F., and J.J.S.S. designed the experiments. T.A.G., J.L., S.H.L., G.S., J.J.S.S., G.H., C.F., R.K.A., N.Y., J.O., K.A.H., and T.E. completed experiments and analyzed data. K.K., M.C.V., and S.Co. performed modeling studies. S.E.H. wrote the manuscript and all authors contributed to editing the manuscript. D.S., S.Ch., S.Co., and S.E.H. supervised experiments and data analyses. S.E.H. obtained funding for the study.

