## Extended Data for "Immune history shapes human antibody responses to H5N1 influenza viruses"

**Extended Tables 1-3**
**Extended Data Figs 1-4**
**Online Methods**
**Extended Data References**

**Table S1. Correlation of antibody titers and influenza subtype imprinting probabilities in healthy individuals.** We computed 95% CIs using Fisher's transformation. Note that the probability of H3N2 imprinting is 1 minus the probability of group 1 imprinting, so correlations with H3N2 and group 1 imprinting are the same but with opposite signs.

| Correlation | Coefficient (95% CI) | P value |
|---|---|---|
| H1 stalk titers - imprinting to H1N1 | 0.43 (0.27, 0.57) | 7.58E-07 |
| H1 stalk titers - imprinting to H3N2 | -0.34 (-0.49, -0.18) | 0.0001 |
| H1 stalk titers - imprinting to group 1 | 0.34 (0.18, 0.49) | 0.0001 |
| H5 stalk titers - imprinting to H1N1 | 0.32 (0.15, 0.48) | 2.87E-04 |
| H5 stalk titers - imprinting to H3N2 | -0.37 (-0.52, -0.21) | 2.76E-05 |
| H5 stalk titers - imprinting to group 1 | 0.37 (0.21, 0.52) | 2.76E-05 |
| H3 stalk titers - imprinting to H1N1 | -0.13 (-0.30, -0.05) | 1.69E-01 |
| H3 stalk titers - imprinting to H3N2 | 0.03 (-0.15, 0.21) | 0.757 |
| H3 stalk titers - imprinting to group 1 | -0.03 (-0.21, 0.15) | 0.757 |
| H7 stalk titers - imprinting to H1N1 | -0.001 (-0.18, 0.18) | 9.87E-01 |
| H7 stalk titers - imprinting to H3N2 | -0.13 (-0.30, 0.05) | 0.158 |
| H7 stalk titers - imprinting to group 1 | 0.13 (-0.05, 0.30) | 0.158 |
| Clade 1 H5 titers - imprinting to H1N1 | 0.39 (0.22, 0.53) | 1.19E-05 |
| Clade 1 H5 titers - imprinting to H3N2 | -0.58 (-0.69, -0.45) | 3.10E-12 |
| Clade 1 H5 titers - imprinting to group 1 | 0.58 (0.45, 0.69) | 3.10E-12 |
| Clade 2.3.4.4b H5 titers - imprinting to H1N1 | 0.35 (0.18, 0.50) | 9.62E-05 |
| Clade 2.3.4.4b H5 titers - imprinting to H3N2 | -0.58 (-0.68, -0.44) | 5.05E-12 |
| Clade 2.3.4.4b H5 titers - imprinting to group 1 | 0.58 (0.44, 0.68) | 5.05E-12 |

**Table S2. Model selection comparing the associations of age, birth year and imprinting probabilities with antibody levels to each antigen. *k* indicates the number of parameters.**

| Antigen | Model | k | loglik | AIC | ∆AIC |
|---|---|---|---|---|---|
| Clade 2.3.4.4b H5 HA | Birth year | 3 | -338.19 | 682.38 | 0.00 |
| | Group 1 imprinting | 3 | -338.35 | 682.71 | 0.33 |
| | Age | 3 | -346.13 | 698.26 | 15.88 |
| | H1N1 imprinting | 3 | -352.26 | 710.53 | 28.15 |
| Clade 1 H5 HA | Birth year | 3 | -378.00 | 762.01 | 0.00 |
| | Group1 imprinting | 3 | -382.62 | 771.24 | 9.23 |
| | age | 3 | -391.09 | 788.19 | 26.18 |
| | H1N1 imprinting | 3 | -393.80 | 793.60 | 31.60 |
| H5 stalk | H1N1 imprinting | 3 | -350.02 | 706.04 | 0.00 |
| | Age | 3 | -350.09 | 706.18 | 0.14 |
| | Birth year | 3 | -350.87 | 707.73 | 1.70 |
| | Group 1 imprinting | 3 | -352.19 | 710.37 | 4.34 |

**Table S3. Bootstrapping analysis comparing how strongly different predictors correlate with titers to each antigen.** Correlation coefficients are shown in absolute value. Boldface values indicate instances where predictor 1 had a stronger correlation with titers than predictor 2 did, based on the 95% bootstrap CI of the difference in correlation coefficients.

| Antigen | Predictor 1 | Predictor 2 | Titer correlation with predictor 1 | Titer correlation with predictor 2 | Observed difference | Bootstrap difference (95% CI) |
|---|---|---|---|---|---|---|
| Clade 2.3.4.4b H5 HA | Birth year | Age | 0.545 | 0.477 | 0.069 | **0.068 (0.034, 0.104)** |
| | Group 1 imprinting | Age | 0.546 | 0.477 | 0.069 | **0.069 (0.001,0.132)** |
| | H1N1 imprinting | Age | 0.374 | 0.477 | -0.103 | -0.103 (-0.210, -0.008) |
| | Group 1 imprinting | Birth year | 0.546 | 0.545 | 0.000 | 0.001 (-0.063, 0.066) |
| Clade 1 H5 HA | Birth year | Age | 0.601 | 0.520 | 0.082 | **0.080 (0.047, 0.113)** |
| | Group 1 imprinting | Age | 0.596 | 0.520 | 0.077 | **0.077 (0.017,0.136)** |
| | H1N1 imprinting | Age | 0.453 | 0.520 | -0.067 | -0.067 (-0.155,0.024) |
| | Group 1 imprinting | Birth year | 0.596 | 0.601 | -0.005 | -0.005 (-0.057,0.049) |
| H5 stalk | Birth year | Age | 0.409 | 0.405 | 0.004 | 0.003 (-0.033,0.043) |
| | Group 1 imprinting | Age | 0.420 | 0.405 | 0.015 | 0.014 (-0.052,0.082) |
| | H1N1 imprinting | Age | 0.361 | 0.405 | -0.044 | -0.040 (-0.143,0.061) |
| | Group 1 imprinting | Birth year | 0.420 | 0.409 | 0.010 | 0.010 (-0.052,0.070) |

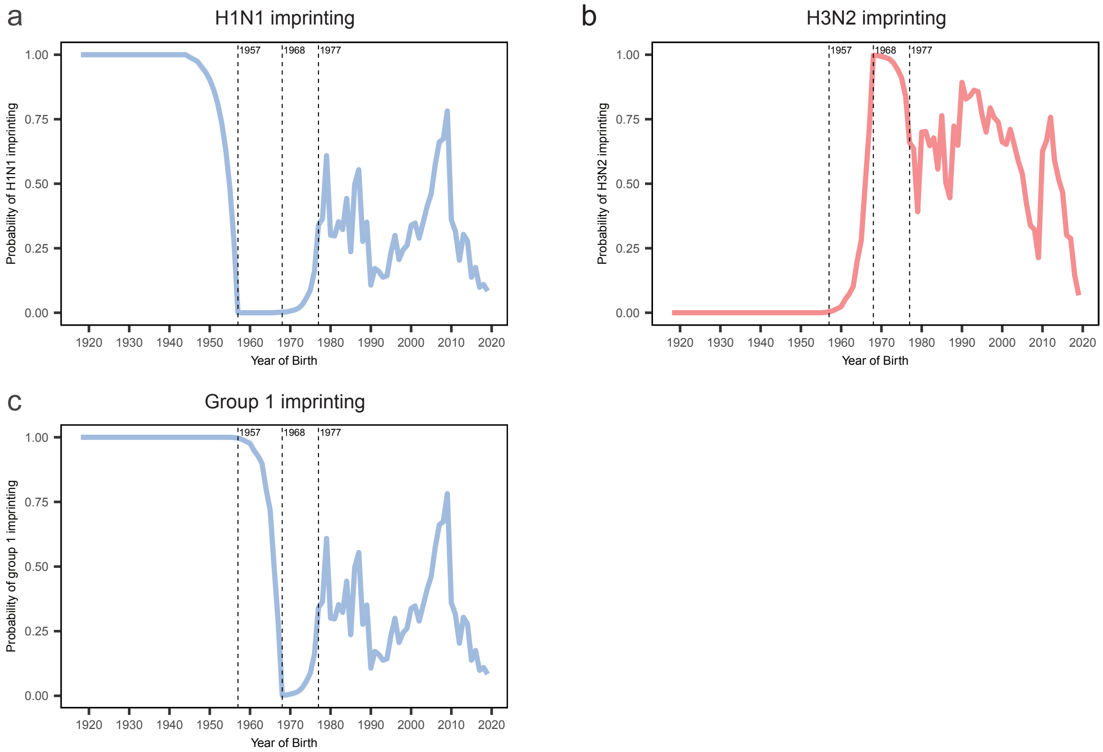


**Extended Data Fig. 1. Imprinting probabilities of influenza subtypes by year of birth.**

Probabilities that an individual was 1st exposed (i.e. ‘imprinted’) in childhood with an (**a**) H1N1 virus, an (**b**) H3N2 virus, or (**c**) group 1 viruses (H1N1 or H2N2) based on their year of birth. Birth year-specific probabilities of immune imprinting to different influenza subtypes were generated using the methods described in Gostic et al.[1] and the *imprinting* R package[2].

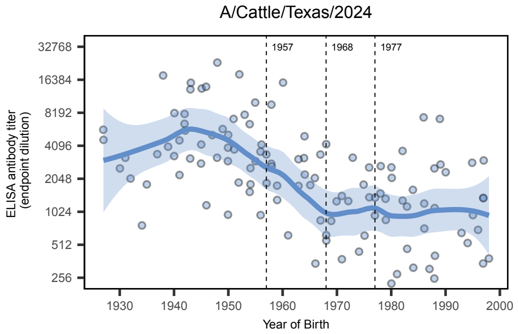


**Extended Data Fig. 2. Group 1 immune imprinting primes antibodies that cross-react to 2.3.4.4b H5N1 isolate currently circulating in cattle.** Sera samples were collected from healthy donors (n=121) at the Hospital of the University of Pennsylvania in 2017 and we quantified IgG binding to clade 2.3.4.4 A/Dairy Cattle/Texas/24-008749-002-v/2024 full length H5 HA. Vertical dashed lines mark years of the 1957 H2N2 and 1968 H3N2 pandemics and 1977 reemergence of H1N1. Each circle represents a geometric mean antibody titer in serum from a single donor from two independent replicates. Trend lines are locally estimated scatterplot smoothing (LOESS) curves (smoothing parameter = 0.4, degree = 2) with 95% confidence intervals.

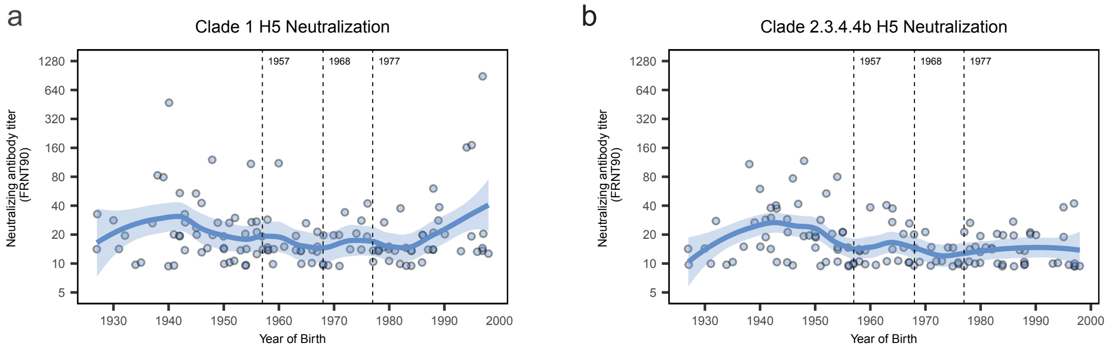


**Extended Data Fig. 3. Most humans possess low levels of neutralizing antibodies against clade 1 and clade 2.3.4.4b H5N1**. Sera samples were collected from healthy donors (n=121) at the Hospital of the University of Pennsylvania in 2017 and we performed neutralization assays using conditionally replicative viruses expressing the HAs from (**a**) clade 1 A/Vietnam/1203/2004 and (**b**) clade 2.3.4.4b A/Pheasant/New York/22-009066-011/2022. Neutralizing titers are reported as reciprocal dilutions of serum that inhibit 90% of virus infection (FRNT90). Vertical dashed lines mark years of the 1957 H2N2 and 1968 H3N2 pandemics and 1977 reemergence of H1N1. Each circle represents a geometric mean antibody titer in serum from a single donor from two independent replicates.  Trend lines are locally estimated scatterplot smoothing (LOESS) curves (smoothing parameter = 0.4, degree = 2) with 95% confidence intervals.

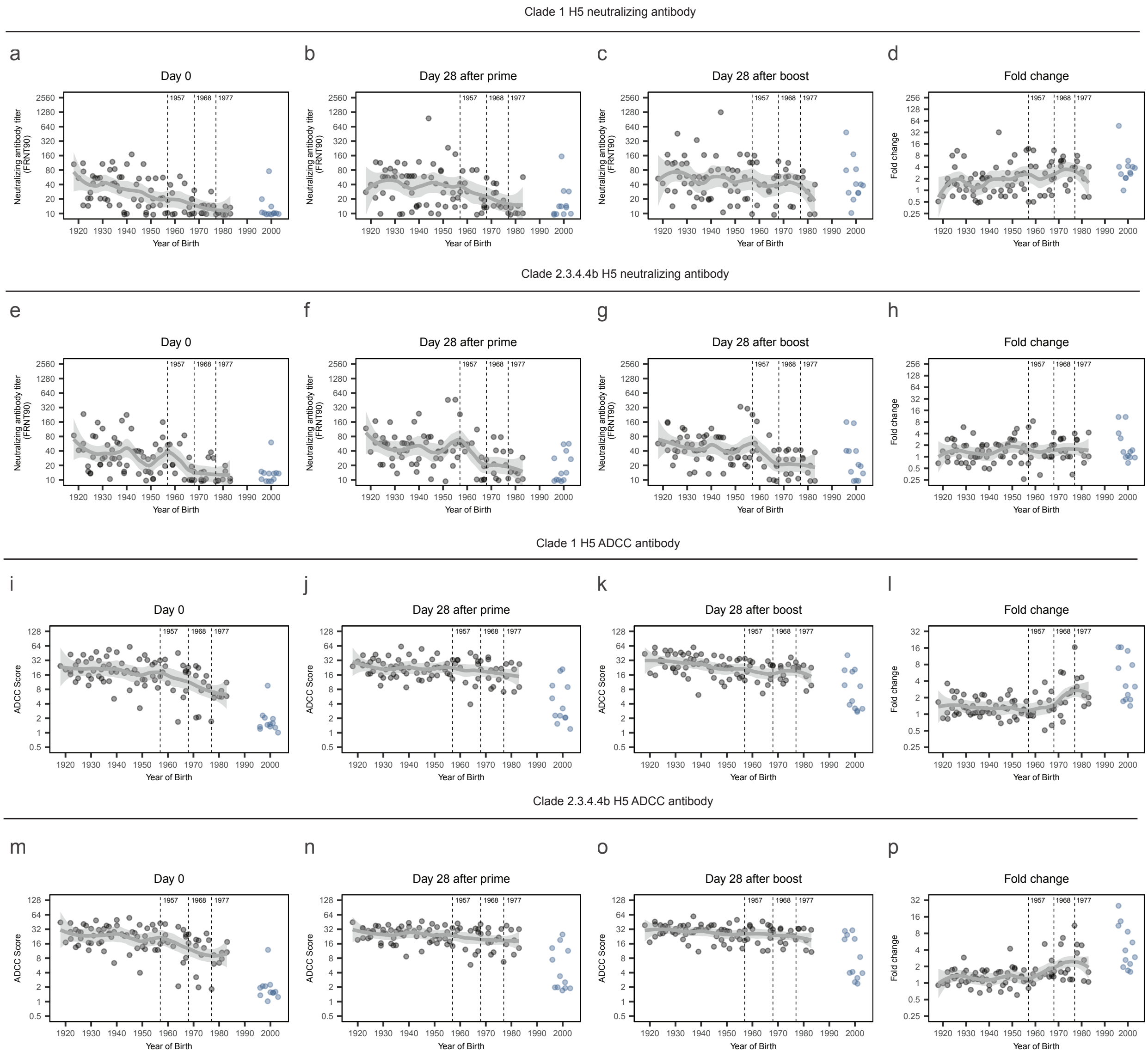


**Extended Data Fig. 4. H5N1 vaccination elicits antibodies that efficiently neutralize vaccine virus and mediate ADCC to matched and mismatched H5N1 viruses.**

Sera samples were obtained from participants (n=100) before (day 0) and 28 days after receiving a first and second dose of an unadjuvanted A/Vietnam/1203/2004 H5N1 vaccine in 2005-2006. We quantified levels of antibodies that neutralized virus with a clade 1 A/Vietnam/1203/2004 H5 HA (**a-d**) and clade 2.3.4.4b A/Pheasant/New York/22-009066-011/2022 H5 HA (**e-h**). Neutralizing titers are reported as reciprocal dilutions of serum that inhibit 90% of virus infection (FRNT90). We quantified levels of antibodies that mediated antibody-dependent cellular cytotoxicity (ADCC) with HAs from a clade 1 A/Vietnam/1203/2004 H5N1 virus (**i-l**) and a clade 2.3.4.4b A/Pheasant/New York/22-009066-011/2022 H5N1 virus (**m-p**). Fold change in panels **d**, **h**, **l**, and **p** were calculated by dividing day 28 post-boost ADCC values by day 0 ADCC values. Samples that did not have neutralizing antibodies were assigned a value of 10. Each circle represents a geometric mean antibody titer in serum from a single donor from two independent replicates. Samples from adults are grey and children are blue. Trend lines are locally estimated scatterplot smoothing (LOESS) curves (smoothing parameter = 0.4, degree = 2) with 95% confidence intervals, fitted to adult samples.

### **Online Methods**

*Human sera*

Donors born between the years of 1927-1998 were recruited to the Hospital of the University of Pennsylvania during the summer of 2017 for blood/serum collection. Additional samples from three clinical trials testing an influenza A (H5N1) vaccine were obtained from the National Institutes of Health (ClinicalTrials.gov #s: NCT00133536, NCT00230750, NCT00115986[3]). Participants received a 45 µg dose of an unadjuvanted A/Vietnam/1203/2004 vaccine and then received a 45 µg dose of the same vaccine 28 days later. We tested sera collected before and 28 days after each vaccination. Informed consent was obtained for all individuals. Experiments using human sera were conducted with the approval of the University of Pennsylvania Institutional Review Boards. Serological experiments were completed using deidentified samples.

*Plasmids*

Expression plasmids were constructed by synthesizing the extracellular domain of A/Vietnam/1203/2004 H5 HA, A/Pheasant/New York/22-009066-001/2022 H5 HA, A/Dairy Cattle/Texas/24-008749-002-v/2024 H5 HA, and A/Shanghai/02/2013 H7 HA, and C-terminally fusing it to the trimeric FoldOn of T4 fibritin, the Avitag sequence, and a hexahistidine affinity tag[4]. These gene fragments were then cloned into the pCMV Sport6 vector, which utilizes a cytomegalovirus (CMV) promoter for mammalian expression.

'Headless' HA expression plasmids encoding recombinant headless HA stalk from either A/California/4/2009 H1N1[5], A/Shanghai/02/2013 H7N9[6], or A/Finland/486/2004 H3N2[6] were provided by Adrian McDermott and Barney Graham from the Vaccine Research Center at the National Institutes of Health. Additional mutations were introduced into 'headless' H1 HA to generate the 'headless' HA from A/Vietnam/1203/2004 H5N1. Secreted BirA-Flag was a gift from Gavin Wright (Addgene plasmid #64395).

*HA protein expression and ‘headless’ HA biotinylation*

Suspension 293F cells (Thermo) grown in FreeStyle medium (Thermo) in flasks were diluted to 1 x $10^6$ cells/mL and HA expression plasmid and plasmid encoding cell surfaced expressed A/Puerto Rico/8/1934 neuraminidase (NA) were transfected with 293fectin reagent into 293F cells (Thermo) according to the manufacturer’s instructions. After 4 days, cells were clarified by centrifugation at 4000 x g for 30 minutes. The supernatant was applied to 5 mL polypropylene columns (Qiagen) containing nickel-nitrilotriacetic acid agarose (Qiagen). Imidazole washing buffer (50 mM $Na_2HCO_3$, 300 mM NaCl, and 20 mM imidazole at pH 8) was applied to the column three times and then HA protein was eluted using imidazole elution buffer (50 mM $Na_2HCO_3$, 300 mM NaCl, and 300 mM imidazole pH 8). Purified HA protein was buffer exchanged into phosphate-buffered saline (PBS) using an Amicon Centrifugal Filter Unit with Ultracel-30 membrane (Millipore). Purified HA proteins were stored at -80°C. Purified ‘headless’ HA proteins were biotinylated *in vitro* using the Avidity BirA-500 kit (product number BirA500; Avidity) or *in situ* by co-transfecting a plasmid encoding a secreted form of the BirA enzyme and also stored at -80°C.[7]

*ELISAs*

ELISAs were performed on 96-well Immulon 4HBX extra high binding flat-bottom plates (Thermo Fisher). For most headless HA ELISAs, 5 µg/mL of streptavidin from *Streptomyces avidinii* (Sigma) was added to each well and plates were incubated at 37°C overnight. After overnight incubation, biotinylated headless HA was added to each well at 1 µg/mL in DPBS (Corning) for 1 hour at room temperature. For full-length HA ELISAs, plates were coated with HA protein at 2 µg/mL in DPBS at 4°C overnight. Following antigen coating, plates were blocked for 1 hour at room temperature either using biotinylation ELISA buffer containing 1X TBS (Bio

Rad), 0.05% Tween 20 (Bio Rad) and 1% bovine serum albumin (Sigma) or standard ELISA buffer containing 1X PBS (Corning), 0.1% Tween 20 (fisher bioreagents), 3% goat serum (Gibco) and 0.5% milk (dot scientific inc). Serum samples were serially diluted two-fold in biotinylation ELISA buffer or standard ELISA buffer, transferred to the ELISA plates, and incubated for 2 hours at room temperature. As a control, HA stalk-specific human mAbs were serially diluted two-fold and added to each plate. This control served to verify equal coating of plates and to determine relative serum titers. Peroxidase-conjugated goat anti-human IgG (product number 109-036-098; Jackson Immunoresearch) was added to all plates at a concentration of 1:5000 in biotinylation ELISA buffer or standard ELISA buffer for 1 hour at room temperature. All plates were developed by adding SureBlue TMB peroxidase substrate (product number 5120-0077; KPL) for 5 minutes at room temperature followed by stopping the reaction with 250 mM HCl solution. The absorbance was measured at 450 nm using a SpectraMax 190 plate reader (Molecular Devices). Plate washing was performed in between each ELISA step by adding and aspirating with 1X PBS (Roche) and 0.1% Tween 20 (Bio Rad) three times using a BioTek 405 LS microplate washer. The nonlinear regression of each serum samples binding was determined using GraphPad Prism. Relative titers were calculated using a consistent concentration of control mAb for each plate and reported as the inverse serum dilution that generated an equivalent optical density (OD) based on the nonlinear regression. The representative Ab titer for each serum sample is the average of two ELISAs performed independently.

*Neutralization assays*

We performed neutralization assays using conditionally replicative influenza viruses that encode GFP instead of PB1, as previously described[8]. Conditionally replicative viruses were rescued and assayed in PB1 complementing cell lines, 293T-CMV-PB1 and MDCK-SIAT1-CMV-PB1-TMPRSS2 cells which constitutively express WSN PB1 protein. The MDCK-SIAT1-

CMV-PB1-TMPRSS2 cells also constitutively express the TMPRSS2 protease, which cleaves and activates HA in the human airways and can substitute for the use of trypsin during influenza infection in vitro[9].

The conditionally replicative viruses were generated from transfection of a co-culture of 4 x $10^4$ 293T-CMV-PB1 and 4 x $10^5$ MDCK-SIAT1-CMV-PB1-TMPRSS2 cells with 350 ng of five bidirectional reverse genetics plasmids encoding the PB2, PA, NP, M, and NS segments from A/Puerto Rico/8/1934, the reverse genetics plasmid pHH-PB1flank-GFP, and the HA from either A/Vietnam/1203/2004 (H5N1) or A/Pheasant/New York/22-009066-001/2022 (H5N1) [10]. We included the corresponding NA for the A/Vietnam/1203/2004 conditionally replicative virus and an N8 from A/Astrakhan/3212/2020 for the A/Pheasant/New York/22-009066-001/2022 conditionally replicative virus since we did not have the corresponding A/Pheasant/New York/22-009066-001/2022 N1 at time of rescue. At 20 hours post-transfection, the media was replaced with neutralization assay media (NAM). NAM has low GFP autofluorescence and contains Medium 199 supplemented with 0.01% heat inactivated FBS, 0.3% BSA, 100 U of penicillin/mL, 100 µg of streptomycin/mL, 100 µg of calcium chloride/mL, and 25 mM HEPES. At 72 hours post-transfection, the supernatants were harvested and cells clarified by centrifugation at 2,000 x g followed by freezing at -80°C. Transfection supernatants were then used to expand the conditionally replicative virus stocks on MDCK-SIAT1-CMV-PB1-TMPRSS2 cells.

The conditionally replicative viruses were normalized by setting up a modified $TCID_{50}$ assay with GFP as a readout. For the modified $TCID_{50}$, conditionally replicative viruses were serially diluted in triplicate across a 96-well plate using NAM. Plates were incubated at 37°C for 1 hour before adding 2.5-8x$10^4$ MDCK-SIAT1-CMV-PB1-TMPRSS2 cells per well at a total volume of 160 µL. Plates were incubated for 40 hours at 37°C followed by measurement of GFP fluorescence intensity on an EnVision multi-mode microplate reader using an excitation wavelength of 485 nm and an emission wavelength of 515 nm. The appropriate conditionally replicative virus dilution to use in the assay was selected based on equal GFP fluorescence intensity at a dilution where the virus starts to linearly reduce in GFP as the virus is diluted (before saturation).

For neutralization assays, sera were pretreated with receptor-destroying enzyme (RDE; Denka Seiken) and heat treated, according to WHO recommended protocols[11]. RDE-treated sera were serially diluted two-fold in NAM across a 96-well plate. Based on the normalization assay, the conditionally replicative viruses were diluted in NAM and added to each well. Plates were incubated for 1 hour before 2.5-8 x $10^4$ MDCK-SIAT1-CMV-PB1-TMPRSS2 cells per well were added. In one row of each plate, no sera were added to measure maximum fluorescence without neutralization, or virus input fluorescence. In a different row on each plate, viruses and sera were not added to measure the background fluorescence of the MDCK-SIAT1-CMV-PB1-TMPRSS2 cells. After 40 hours of incubation at 37°C, GFP fluorescence intensity was measured. Percent infectivity was calculated by dividing the background-subtracted fluorescence of the serum containing wells by the background-subtracted virus input fluorescence. Percent neutralization was determined by subtracting the percent infectivity from 100. Neutralization titers were expressed as the inverse of the highest dilution that inhibited 90% of GFP virus infection (FRNT90). The representative neutralization titer for each serum sample is the average of two neutralization assays performed independently.

*Jurkat/Fc Receptor Reporter ADCC assays*

To prepare antigen presenting cells, HEK293T/17 cells (ATCC, CRL-11268) were transfected with plasmid DNAs to express membrane bound A/Vietnam/1203/2004 or A/Pheasant/New York/22-009066-001/2022 HA proteins. Twenty-four hours post-transfection, transfected cells were dissociated, and HA expression confirmed by flow cytometry after staining cells with the pan-stalk, cross-reactive mAb CR9114. Three thousand HEK293T17 expressing

A/Vietnam/1203/2004 or A/Pheasant/New York/22-009066-001/2022 HA were plated in 384 well assay plates (Corning 3765) in 20 µL of DMEM supplemented with 10% fetal bovine serum. Twenty-four hours after plating, 10 µL of growth medium was robotically aspirated from assay plates. An 8-point dilution series (4-fold dilution between concentrations) of each serum sample was robotically prepared by diluting samples in 2% (w/v) BSA in PBS. 2 µL of the 8-point dilution series of patient serum was robotically transferred to assay plates, yielding a final serum dilution range of 1:100 to 1:1,600,000. A dilution series of control mAbs CR9114, FI6, 70-1F02, EM-4C04, ranging from 1 µg/mL to 0.00046 µg/mL, and the negative control (2% (w/v) BSA in PBS, n=16) was included on each assay plate. Nine thousand Jurkat/FCGR3A-V158/NFAT-Luciferase reporter cells in 10 µL of RPMI supplemented with 10% ultra low-IgG fetal bovine serum was added to assay plates. Assay plates were incubated at 37°C/5% $CO_2$ for 5 hours.

Assay plates were removed from the incubator for 1 hour to equilibrate to room temperature prior to adding 20 µL of Brite-lite Luciferase Reagent (revvity). Luminescence was measured on an EnVision Xcite Multilabel Plate Reader (revvity), using the ultrasensitive luminescence measurement technology. Assay well data were normalized to aggregated negative control wells and expressed as antibody-dependent signal-to-background (S:B = sample well/$NegC_{Avg}$). ADCC scores were defined as the S:B at a 1:1600 dilution of the patient serum.

*Imprinting probabilities*

We estimated imprinting probabilities for people born in the United States using the *imprinting* package with default parameters, which implements the approach from Gostic et al.[1,2]. Briefly, the method assumes a constant annual probability of exposure to influenza and uses historical subtype frequencies to calculate the probability that a person born in a particular year was first infected with a subtype or group of interest.

*Statistical analyses*

For robustness, we used two different statistical approaches to test if antibody titers were associated more strongly with birth year or imprinting probabilities than with age. First, we fitted separate linear regressions with titers to a particular antigen as the response value and either age, birth year, probability of group 1 imprinting or probability of H1N1 imprinting as the predictor value. We compared the models using the Akaike Information Criterion (AIC). Second, we compared pairs of predictors by taking the absolute value of the Spearman correlation coefficient between each predictor and antibody titers, computing the difference between those values, and constructing a 95% confidence interval for that difference using bootstrapping (i.e., repeating the calculations after resampling the observations with replacement 1,000 times). A 95% CI that does not overlap zero indicates that the predictors differ in how strongly they correlate with antibody titers. Code implementing the analyses is available at https://github.com/cobeylab/H5_titers_vs_imprinting

*Data availability*

All data are included in the manuscript and there are no restrictions. The code implementing the analyses is available at https://github.com/cobeylab/H5_titers_vs_imprinting

…

### Extended Data References


1. Gostic, K.M., Ambrose, M., Worobey, M. & Lloyd-Smith, J.O. Potent protection against H5N1 and H7N9 influenza via childhood hemagglutinin imprinting. *Science* **354**, 722-726 (2016).
2. Gostic, K. imprinting: Calculate Birth Year-Specific Probabilities of Immune Imprinting to Influenza. R package version 0.1.0. https://cran.r-project.org/web/packages/imprinting/index.html (2022).
3. Treanor, J.J., Campbell, J.D., Zangwill, K.M., Rowe, T. & Wolff, M. Safety and immunogenicity of an inactivated subvirion influenza A (H5N1) vaccine. *The New England journal of medicine* **354**, 1343-1351 (2006).
4. Whittle, J.R.*, et al.* Flow cytometry reveals that H5N1 vaccination elicits cross-reactive stem-directed antibodies from multiple Ig heavy-chain lineages. *Journal of virology* **88**, 4047-4057 (2014).
5. Yassine, H.M.*, et al.* Use of Hemagglutinin Stem Probes Demonstrate Prevalence of Broadly Reactive Group 1 Influenza Antibodies in Human Sera. *Scientific reports* **8**, 8628 (2018).
6. Corbett, K.S.*, et al.* Design of Nanoparticulate Group 2 Influenza Virus Hemagglutinin Stem Antigens That Activate Unmutated Ancestor B Cell Receptors of Broadly Neutralizing Antibody Lineages. *mBio* **10**(2019).
7. Bushell, K.M., Sollner, C., Schuster-Boeckler, B., Bateman, A. & Wright, G.J. Large-scale screening for novel low-affinity extracellular protein interactions. *Genome Res* **18**, 622-630 (2008).
8. Doud, M.B., Hensley, S.E. & Bloom, J.D. Complete mapping of viral escape from neutralizing antibodies. *PLoS pathogens* **13**, e1006271 (2017).
9. Lee, J.M.*, et al.* Deep mutational scanning of hemagglutinin helps predict evolutionary fates of human H3N2 influenza variants. *Proceedings of the National Academy of Sciences of the United States of America* **115**, E8276-E8285 (2018).
10. Bloom, J.D., Gong, L.I. & Baltimore, D. Permissive secondary mutations enable the evolution of influenza oseltamivir resistance. *Science* **328**, 1272-1275 (2010).
11. World Health Organization. *Manual for the laboratory diagnosis and virological surveillance of influenza*, (World Health Organization, Geneva, 2011).
